# A Mathematical Model Approach for Prevention and Intervention Measures of the COVID–19 Pandemic in Uganda

**DOI:** 10.1101/2020.05.08.20095067

**Authors:** Fulgensia Kamugisha Mbabazi, Yahaya Gavamukulya, Richard Awichi, Peter Olupot–Olupot, Samson Rwahwire, Saphina Biira, Livingstone S. Luboobi

**Affiliations:** Department of Mathematics, Faculty of Science and Education, Busitema University, Tororo, Uganda; Department of Natural Sciences, Uganda Martyrs’ University, Kampala, Uganda; Department of Biochemistry and Molecular Biology, Faculty of Health Sciences, Busitema University, Mbale, Uganda; Department of Public Health, Faculty of Health Sciences, Busitema University, Tororo, Uganda; Mbale Clinical Research Institute, Mbale, Uganda; Department of Polymer, Textile and Industrial Engineering, Faculty of Engineering, Busitema University, Tororo, Uganda; Department of Physics, Faculty of Science and Education, Busitema University, Tororo, Uganda; Institute of Mathematical Sciences, Strathmore University, Nairobi, Kenya

**Keywords:** COVID-19, SEIR model, Awareness, Infection rate, control measures

## Abstract

The human–infecting corona virus disease (COVID–19) caused by the novel severe acute respiratory syndrome corona virus 2 (SARS–CoV–2) was declared a global pandemic on March 11th, 2020. Current human deaths due to the infection have raised the threat globally with only 1 African country free of Virus (Lesotho) as of May 6th, 2020. Different countries have adopted different interventions at different stages of the outbreak, with social distancing being the first option while lock down the preferred option for flattening the curve at the peak of the pandemic. Lock down is aimed at adherence to social distancing, preserve the health system and improve survival. We propose a Susceptible–Exposed–Infected–Expected recoveries (SEIR) mathematical model to study the impact of a variety of prevention and control strategies Uganda has applied since the eruption of the pandemic in the country. We analyze the model using available data to find the infection–free, endemic/infection steady states and the basic reproduction number. In addition, a sensitivity analysis done shows that the transmission rate and the rate at which persons acquire the virus, have a positive influence on the basic reproduction number. On other hand the rate of evacuation by rescue ambulance greatly reduces the reproduction number. The results have potential to inform the impact and effect of early strict interventions including lock down in resource limited settings and social distancing.

## Introduction

Corona Virus Disease 2019 (COVID–19), first discovered in Wuhan City, Hubei Province, China on December 31st 2019 [7], has established itself as the most devastating global pandemic todate. The disease has not respected borders, socio–economic developments of countries or states, and personal status. COVID–19 caused by Severe Acute Respiratory Syndrome Corona Virus 2 (SARS–CoV–2) has spread worldwide [1]. Even in countries and states with very high levels of emergency response preparedness, COVID–19 has had early and projected ramifications in all areas of life including health, economy, education, travel, development and security. Globally, the number of cases confirmed to the disease has surpassed 3.5 million people with over 248,313 deaths (CFR: 7%) and 1,157,014 recoveries [5]. The evolution of the current pandemic has dis–proportionally affected various countries. The top most affected countries both in confirmed cases and mortality burden include: USA, Spain, Italy, France, UK and Germany. With USA showing the highest death toll 68,602 persons (with 5.8% Case fatality rate), as of May 4th, 2020 [5]. In Africa, an estimated 44,483 confirmed cases; 1,801 death, with case fatality rate of 4% and 14,921 recoveries have occurred, as of May 4th, 2020 [23]. Countries that have not reported COVID–19 cases as of, May 4th, 2020 include: Kiribati, Lesotho, Marshall Islands, Micronesia, Nauru, North Korea, Palau, Samoa, Solomon Islands, Tonga, Turkmenistan, Tuvalu, Vanuatu [25]. The pandemic however, has both direct and indirect effects and ramifications in all sectors of life including health, economy, trade, travel, education and governance.

In Uganda, the index case was confirmed on March 21, 2020 [4]. Despite immediate lock down and intense public health interventions including contact tracing, the country has registered eighty–nine (89) cases as of May 4th, 2020 [5]. Majority of these cases are imported cases including recent truck drivers from the East African region. The total number of foreign truck drivers who have tested positive for COVID–19 is thirty (30), of these nineteen have returned to their respective countries whereas eleven are admitted at different hospitals in Uganda [6]. The community transmission through contacts has emerged and the extent of which remains unknown since many of those who traveled into the country remain untraced while others are still under self quarantine. Being a landlocked country with porous borders and all countries surrounding it having reported more cases than reported in Uganda (except South Sudan), the risk of the pandemic spreading widely is high. The progress on management of the country outbreak of COVID–19 in Uganda is promising. The Uganda Ministry of Health COVID–19 updates as of May 4th, 2020 indicate that a total of thirty nine thousand, two hundred thirty two (39,232) persons have been tested, of whom eighty nine (89) are confirmed cases, eight hundred sixty one (861) have been discharged from institutional quarantine, four hundred forty six (446) are under institutional quarantine, one thousand three hundred two (1,302) are contacts listed, eight hundred eight (808) are under follow up, 141 are under self quarantine, eighteen (18) are active cases and fifty two (52) have fully recovered following successful treatment [3,6]. The distribution of COVID–19 confirmed cases by residence is shown in Fig 1.

**Fig. 1.**
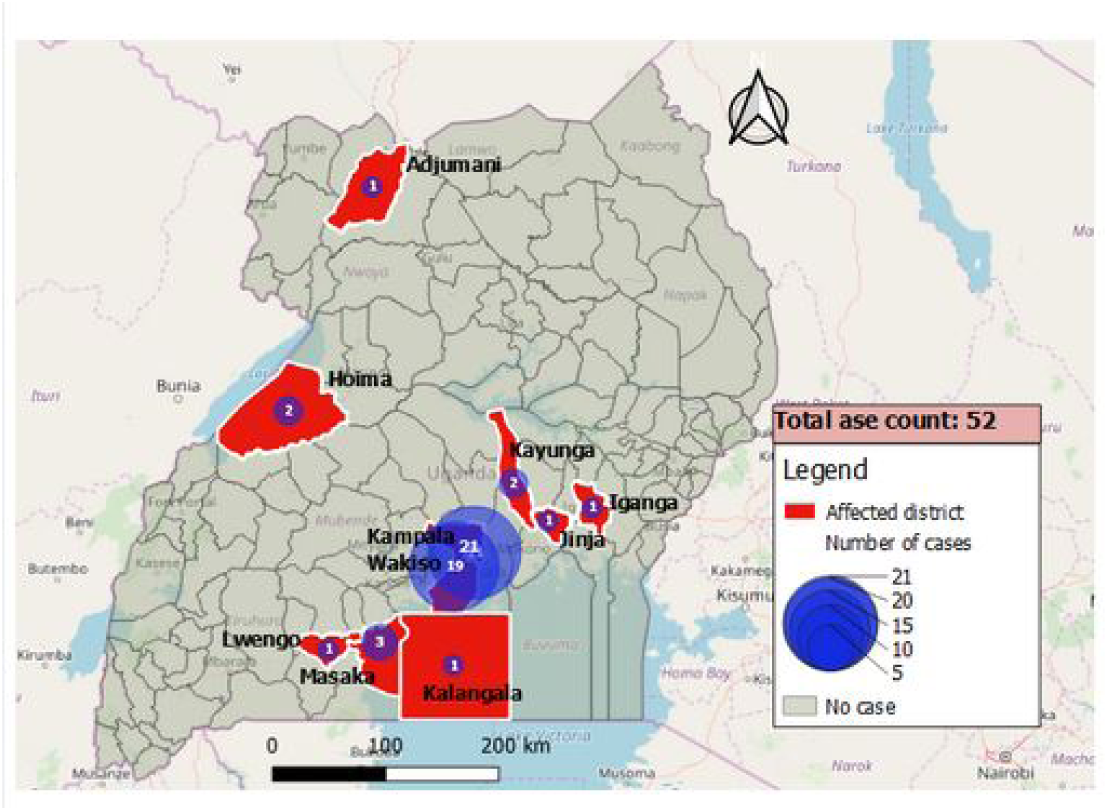
Distribution of COVID—19 confirmed cases by place of residence in Uganda [6].

Like in many countries, especially with limited resources, public health, community engagement and social science intervention were urgently and rapidly rolled out by the Uganda government. To date, emphasis and law enforcement on social distancing, lock down, travel bans and mass sensitizations on the virus prevention methods to control the COVID–19 Pandemic are in place.This is in addition to updates and directives by the President of the Republic of Uganda that are usually given every after two or three days. However, the rural population may remain unaware of the said interventions to prevent and control COVID–19 spread in the communities. This is out of the fact that, many don’t have access to electricity and communication media. Televisions, Radios and Newspapers are not affordable to the majority. This calls for the need to have awareness programs extended to the rural areas in order to educate the population about the spread of the COVID–19 virus, its prevention and control measures. This would limit the number of exposed and infected persons in the country.

The country is implementing a model of early lock down and fractional testing of high risk populations including travelers and contacts of confirmed cases. This is as opposed to models in China, Europe and USA where the lock down followed unprecedented number of cases and deaths. The only similar model of the developed world to that employed by the Uganda government is the Greece model. Most African countries including those in the East African Region have also followed early lock down. The biggest percentage of Uganda population is rural population and may be at the risk of contracting the disease. Awareness by mass media, are limited by existing resources and other socioeconomic factors, and it is generally difficult for these awareness programs to be disseminated to the whole host population.

Mathematical models have been used during the outbreak of COVID–19 in China, Italy and other countries to give direction to policy and decision makers in government institutions. The commonly used model is the SEIR and include works of [12,13,18,19]. A study done by Rovetta [20] has used the SEIR model to predict and inform governments of different countries about the COVID–19 pandemic. The models have been modified by Hang et al. [14], Zhu and Zhu [15], Wan et al [16] and Cao et al. [17] to include the asymptomatic classes, symptomatic classes, quarantine population, self isolation and death classes in order to assess the impact of the disease and predict the epidemic in the populations.

In our model [8] designed for the early in–country outbreak of COVID–19, we predicted how the rate at which COVID–19 would spread in the country without prevention and intervention measures. Approximately one hundred twenty seven (126) persons were predicted to have contracted the disease in two weeks and four thousand, three hundred sixty nine (4,370) persons to have contracted the disease in a month if no prevention and intervention measures are put in place (see Fig 2). We have since developed a hypothesis based on these data and model. We hypothesize that with prevention and intervention measures put in place the trend would change. This hypothesis is informed by our research question that there is a proportion of the community that is not aware of COVID–19 pandemic prevention and intervention control measures in Uganda. This study is therefore set to model the COVID–19 pandemic that incorporates prevention and intervention measures with awareness to reduce the previous projected infected numbers in order to reduce the disease spread and consequently flatten the COVID–19 Pandemic Curve in Uganda. We adopt the vital dynamics of the SEIR that incorporates awareness through media coverage, prevention and control measures.

**Fig. 2.**
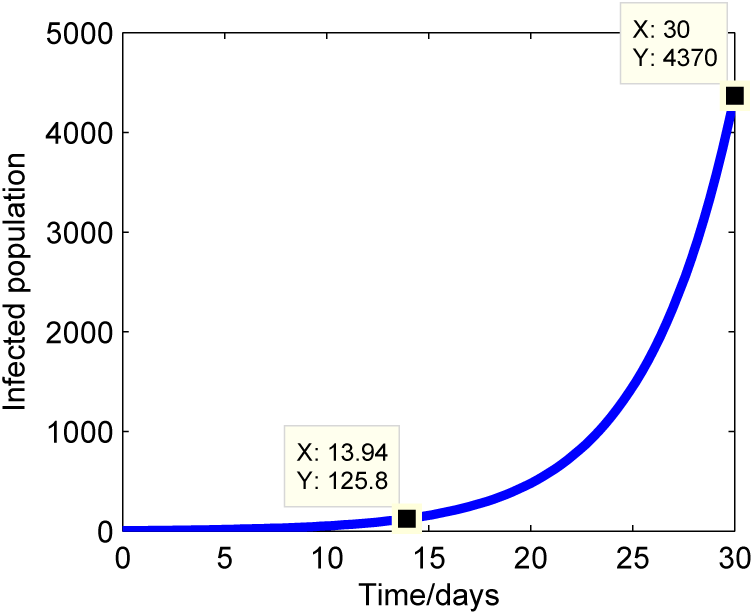
The exponential projection of the number of infected individuals due to COVID—19 in Uganda without interventions (March, 2020)

## METHODOLOGY

### The SEIR model

The model under consideration is a deterministic

”Susceptible–Exposed–Infected–Expected recoveries (SEIR)” compartmental model based on the dynamics of the disease including epidemiological status of individuals and control measures (physical distancing, quarantine, curfew and lock down) currently used in Uganda. The population consists of four compartments with the susceptible sub–population divided into aware persons *S_a_(t)* and unaware persons *S_u_(t)* at time *t*, the exposed/quarantined persons (individuals with a travel history) *E(t)* and infected persons (infectious with disease symptoms) *I*(*t*), the infected persons on treatment expected to recover *R(t)* at time *t*. If the tracing of contact is considered, a fraction p of persons exposed to COVID–19, is quarantined. The quarantined persons can either transfer to the infected compartment or to aware susceptible compartment *S_a_(t)* depending on whether infectious or not.

We assume Uganda to be a closed system with a constant population *N* = 45, 395, 554 during the epidemic and the exposed population initially to consist all returnees *E* = 18,128. The unaware population is increased through aware persons loosing memory about information on prevention and intervention measures. The unaware persons get infected either in contact with infected individuals or with infected objects and transfer to the exposed population at a mass incidence rate *bS_u_I*, where b=*β*(*d* + *q* + *l* + *h*), comprising the product of the transmission rate with the sum of control measures: physical distancing *d*, lock down *l*, quarantine q and hygiene practices *h* put in place. The unaware population get information about COVID–19 from media, implement it at a rate ζ and thus transferring to the aware population.

The aware sub–population is increased by persons who implement the existing prevention and control measures learned from media. The aware population may get in contact with infected objects and persons because human tend to forget due to some social factors and transfer to the exposed population. The exposed population initially consists of persons with a travel history that were checked on arrival, quarantined for fourteen days (incubation period), however some persons never went through the process of checking and mixed with the community. The quarantined persons in the exposed population are tested after the incubation period of 14 days, if tested negative a proportion (1 − *p)δE* transfer to the aware population and a proportion *pδE* transfer to the infected class, with *p* the proportion of persons that acquire infection and *δ* the incubation rate.

The infected population is assumed to decrease at a rate *r* + *q_i_* + *q_s_*, where *r* is the rate of evacuation by rescue ambulances, *q_s_* is the rate at which individuals with mild symptoms isolate themselves from the population, *q_i_* is the rate at which infected individuals are quarantined in institutional centres. Infected individuals transfer to the sub population of expected recoveries at a rate *γ*. The infected population quarantined under government institutions and expecting to recover at time *t* is increased by persons who are found to have symptoms, retained in hospitals for further administration of treatment. Persons who respond to treatment are removed at a rate *ϕ*.

### State variables and model parameters

**Table 1.** Description of state variables and their possible values.

| State variable | Description | Initial value | Source |
| --- | --- | --- | --- |
| $S(t)$ | Number of susceptible persons at time $t$ | 45,427,637 | [21] |
| $S_u(t)$ | Number of unaware persons at time $t$ | 33,752,735 | Estimate |
| $S_a(t)$ | Number of aware persons at time $t$ | 11,674,902 | Estimate |
| $E(t)$ | Number of exposed persons at time $t$ | 18,128 | [6] |
| $I(t)$ | Number of infectious persons at time $t$ | 1 | [3] |
| $R(t)$ | Number of infected persons on treatment expected to recover at time $t$ | 0 | assumed |
| $M(t)$ | Number of Media type that run awareness programs in a given locality at time $t$ | 4 | assumed |

The compartmental diagram of the model is shown in Fig 1.

**Fig. 3.**
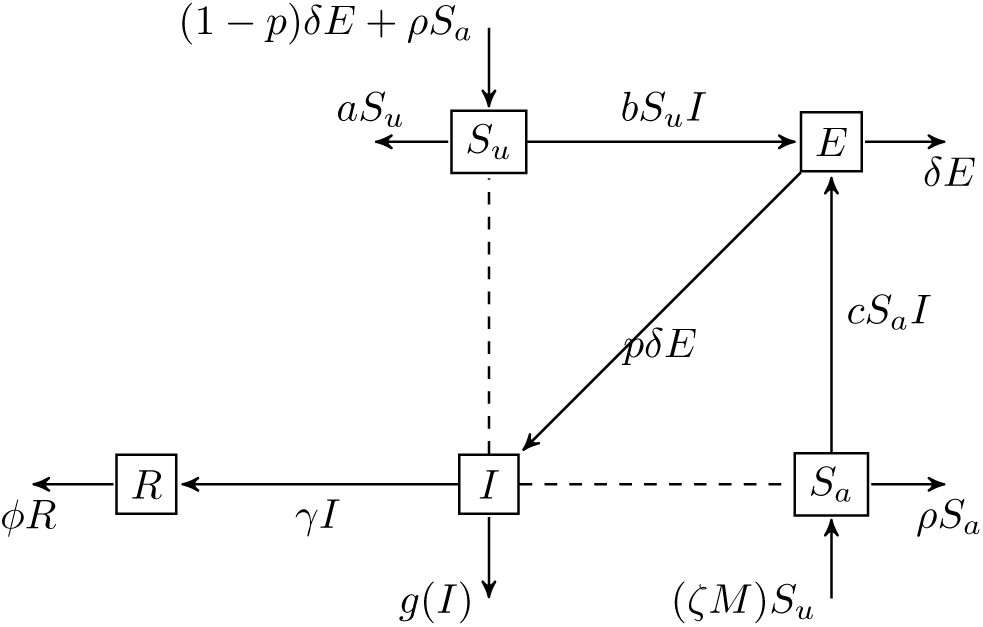
A schematic diagram showing the dynamics of COVID–19 (Corona virus). The dotted lines represent contacts made by individuals in the respective classes and the solid lines show transfer from one class to another.

with

*g(I)* = (*r* + *q_i_* + *q_s_*)*I*,
*a* = *ζM*,
*b* = *β*(*d* + *q* + *l* + *h*),
*c = β(d + k)*.

### Model equations

In this study, upon giving a transition diagram, assumptions and description of model parameters, the ordinary differential equations for the population change of each sub–population are stated as

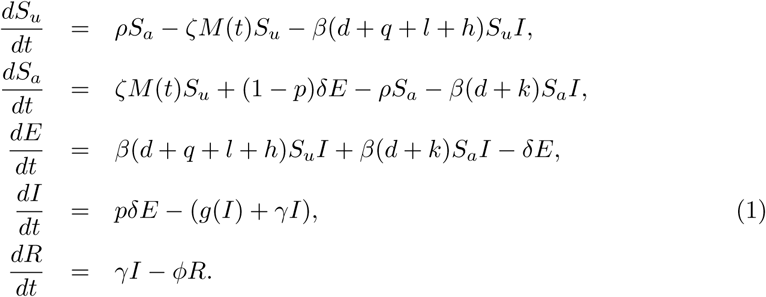

with 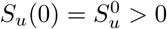, 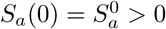, *E*(0) = *E*_0_ ≥ 0, *I*(0) = *I*_0_ ≥ 0, *R*(0) = *R*_0_ ≥ 0, *M*(0) = *M*_0_ ≥ 0, *g(I*) = (*r* + *q_i_* + *q_s_*)*I*.

### Parameter estimation

Basic data used in this study were obtained from the daily epidemic announcements by media and Ministry of Health. Release of cumulative data about COVID–19 in terms of confirmed cases of infected, critical, total deaths, recovered and cumulative tested cases [5]. We assume persons that had a travel history to be quarantined in institutional centers for 14 days. Information from official websites and previous studies done as of April 27, 2020.

The initial population conditions with regard to system (1) for Uganda are set to: *S_u_*(0) = 33, 752, 735, *S_a_*(0) = 11, 674, 902, *E*(0) = 18,128, *I*(0) = 0, *R*(0) = 0. The latency period is assumed to be Erlang distributed with mean 5.2 days (SD 3.7) [11].

We estimate the removal rate of infected persons on treatment expected to recover = 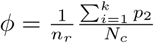, with 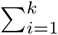 *p*_2_ = cumulative recoveries, *N_c_* = total of confirmed cases, *n_r_* = number of days for which an individual takes to recover, 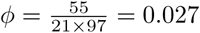.

The average disease duration for COVID–19 is 21 days, hence the rate of recovery is given by 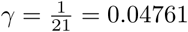 per day.

### The steady states and the reproduction number, *R_n_*_0_

The positive invariant set Ω has two possible steady states, that is the infection free steady state and the endemic steady state.

#### The infection–free steady state

For this state the population is free of the infection at the beginning of the epidemic.

The infection free steady state 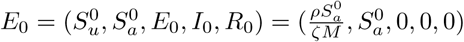 on the set Ω

#### The endemic steady state

The endemic steady state is got by setting the R.H.S of Eq (1) to zero. That is:

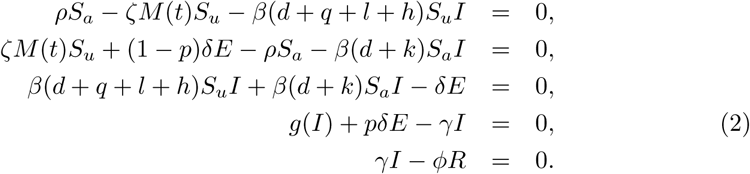

Hence the endemic steady state 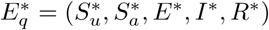 on a set Ω is given as, where

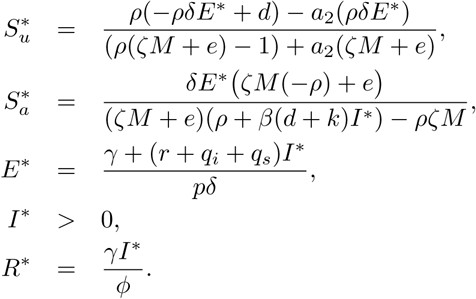

With

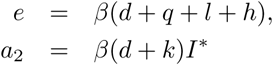

We note that, 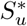 is biologically feasible provided

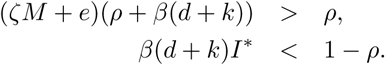

Whereas

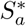 is biologically feasible provided

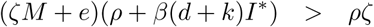

The biological meaning of this endemic state is that the disease establishes itself in the population and persists for a long period.

#### The reproduction number, *R_n_*_0_

The basic reproduction number (*R_n_*_0_) of COVID–19 disease, indicates the transmissibility of the Corona virus, as a representative of the average number of new infections produced by one infectious person in a wholly naive population. If *R_n_*_0_ is less than unity the infection is likely to perish out whereas if *R_n_*_0_ is greater than unity the infection is likely to propagate and persist in the population. The basic reproduction number is a vital threshold in modeling infectious diseases that show the threat of an infectious pathogen with respect to the epidemic spread. The magnitude of *R_n_*_0_ is significant in determining the severity of the disease, and help to draw plans and design control strategies. Since COVID–19 epidemic is in it’s early stage of spread, we assume 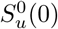, 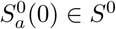 to be near the infection–free steady state value 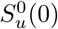, 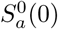, and approximating differential equations of the exposed and infected classes to a linear system:

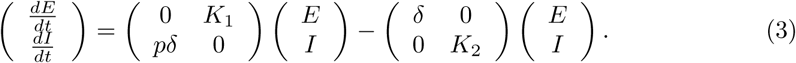

with 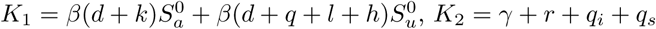

Eq (3), the linearization has been separated into two parts, i.e. first matrix represents infection rates and the second matrix represents a combination of transition rates and growth rate.

Let *G* =the matrix of infection rates and *U* =the matrix combination of transition rates and growth rate.

Such that

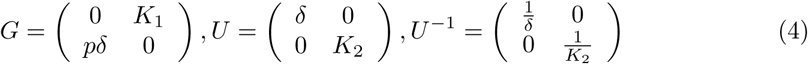

We find the next generation matrix *GU*^−1^. Then the spectral radius of the product of *G* and *U*^−1^ to be the reproduction number [9].

From Eqs (4).

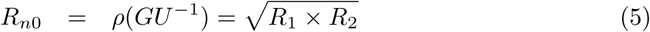

Eq (5) yields an interesting reproduction number *R_n_*_0_ of a geometric mean with two terms *R*_1_ and *R*_2_ with:

R_1_ = *p* (the rate of acquiring infection) and 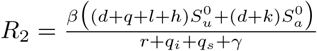 (Infection rate for aware and unaware persons/Sum of output rates).

Using initial parameter values in Table 2, the reproduction number is estimated to be *R_n_*_0_ = 0.468. The reason of this geometric mean during the early stages of COVID–19 spread is because COVID–19 has the potential to re–emerge in the population upon successful eradication.

**Table 2.**
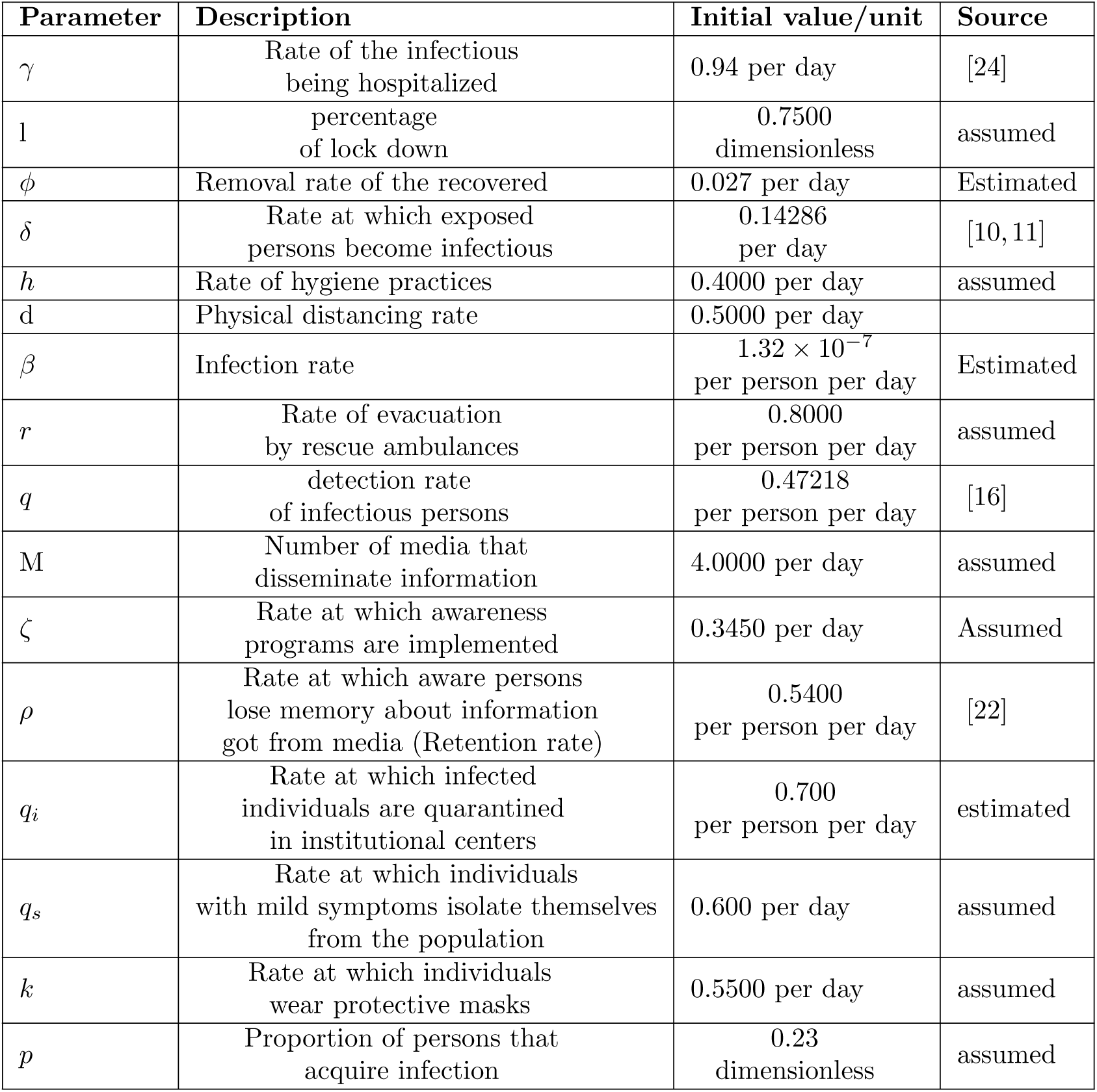
Description of parameters and their possible values.

## Sensitivity analysis and Numerical results

### Sensitivity analysis of model parameters with respect to *R*_0_

In the fight of an emerging disease, the Ugandan government has taken several strategies to prevent and control the disease spread. The prevention and control measures include: social distancing, hand washing, wearing of masks, lock down, travel bans, curfew and media sensitizations. To scrutinize and assess the potential effectiveness of these strategies, we do the sensitivity analysis for the dynamic model parameters on the threshold number *R_n_*_0_, which gives a best measure for the control of an epidemic in the population. The sensitivity analysis establishes the significance of every parameter to disease spread.

Definition: The elasticity / normalized forward sensitivity index measures the relative change of *R_n_*_0_ with respect to a parameter *z*, defined by

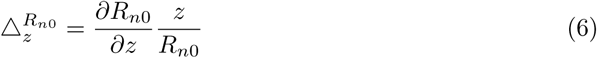

The sign of elasticity index explains whether *R_n_*_0_ increases (positive sign) or reduces (negative sign) with the parameter while the magnitude establishes the relative significance of the parameter (see Table 3). Such indices give direction to decision /policy makers on important parameters to be targeted for cost effectiveness and practical control strategies.

**Table 3.** Sensitivity indices (S.I) of *R_n_*_0_ with respect to the model parameters.

| Code | Parameter | Sensitivity index |
| --- | --- | --- |
| 1 | $\beta$ | +1.0000 |
| 2 | $p$ | +1.0000 |
| 3 | $d$ | +0.3699 |
| 4 | $k$ | +0.2925 |
| 5 | $l$ | +0.11560 |
| 6 | $h$ | +0.0832 |
| 7 | $\gamma$ | 0.0000 |
| 8 | $q_s$ | -0.2790 |
| 9 | $q_i$ | -0.3259 |
| 10 | $r$ | -0.3725 |

Fig 4 indicate the Infection rate *β* and proportion of persons that acquire infection *p* to have a stronger impact on the threshold number *R_n_*_0_ than other parameters. When the said parameters are increased by 10%, the basic reproduction number increases from *R_n_*_0_ = 0.468 to *R_n_*_0_ = 1.4569 which is slightly more than thrice the original value. This implies that the population is likely to have more infected cases that may require stringent measures to be put in place. In addition the rate of evacuation of infected individuals from the community by the rescue ambulances *r*, significantly reduces *R_n_*_0_ = 0.468 to *R_n_*_0_ = 0.2539 by 46%. This implies that the faster infected persons are taken to hospital the lesser the number of infected individuals in the community. On the other hand the rate at which individuals with mild symptoms isolate themselves from the population *q_s_*, the one with less sensitivity index, once increased by 10% there is a reduction in the basic reproduction number from *R_n_*_0_ = 0.468 to *R_n_*_0_ = 0.279 which is a 40% decrease. This implies that the rate at which persons quarantine themselves would reduce the disease spread in the population. The Rate of the infectious being hospitalized *γ* shows no change in the basic reproduction number.

**Fig. 4.**
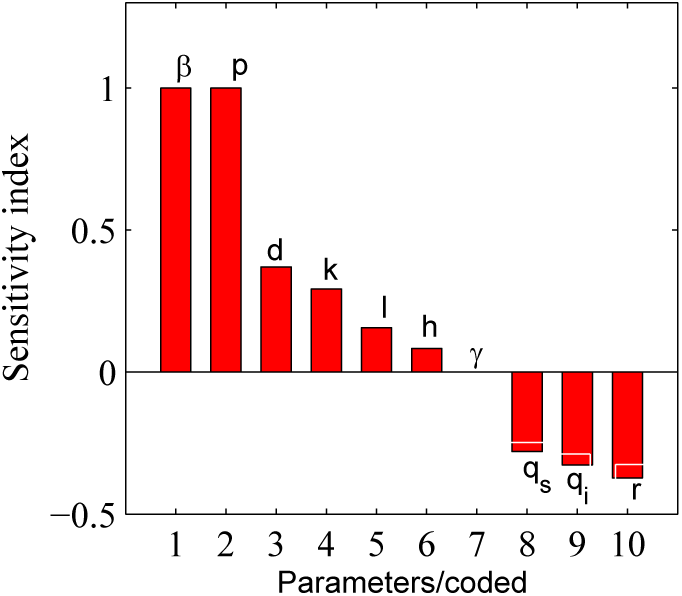
Sensitivity analysis of different parameters on the basic reproduction number *R_n_*_0_.

## Numerical results and discussion

We apply the Runge Kutta fourth and fifth order to solve system (1) with the help of MATLAB.

Fig 5 shows the epidemic curve for the infected and the expected recovered persons attaining peak points (turning points) at (52 days, 177 persons) and (158 days, 1679 persons) respectively.

**Fig. 5.**
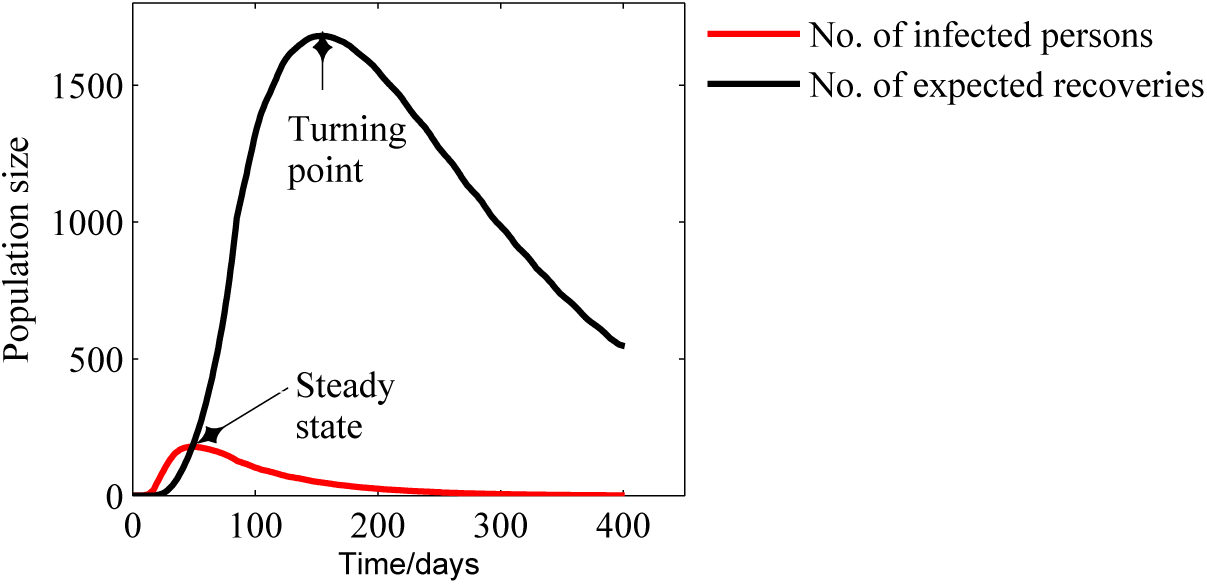
Model (1) simulations for time series projection for the infected and expected recovered persons from COVID—19 in the presence of prevention and control measures.

A unique intersection point called the steady state point at which equal numbers of infected and expected recovered persons are equalized is (51 days, 178 persons). The sudden decrease in the number of infected persons is a resilient confirmation for the effectiveness of the available facilities in the health care systems. Whereas an increase of the number of expected recoveries is evidence of enough facilities available. The disease slows down and health institutions cope with the sick by giving them attention. The government has the potential to eradicate the disease in fourteen months (1 yr and 2 months).

From our model system (1) Fig 6 (a) that is generated, we project the infected population to reach a peak time in one month and 22 days with an estimated 177 infected persons. The government of Uganda has done her best to prevent and control the disease to see that no death has been confirmed as per May 7, 2020. However, Fig 6(b) shows the likelihood of COVID–19 disease re–emerging in the population after 430 days ≈ one (1) year and (10) ten days.

**Fig. 6.**
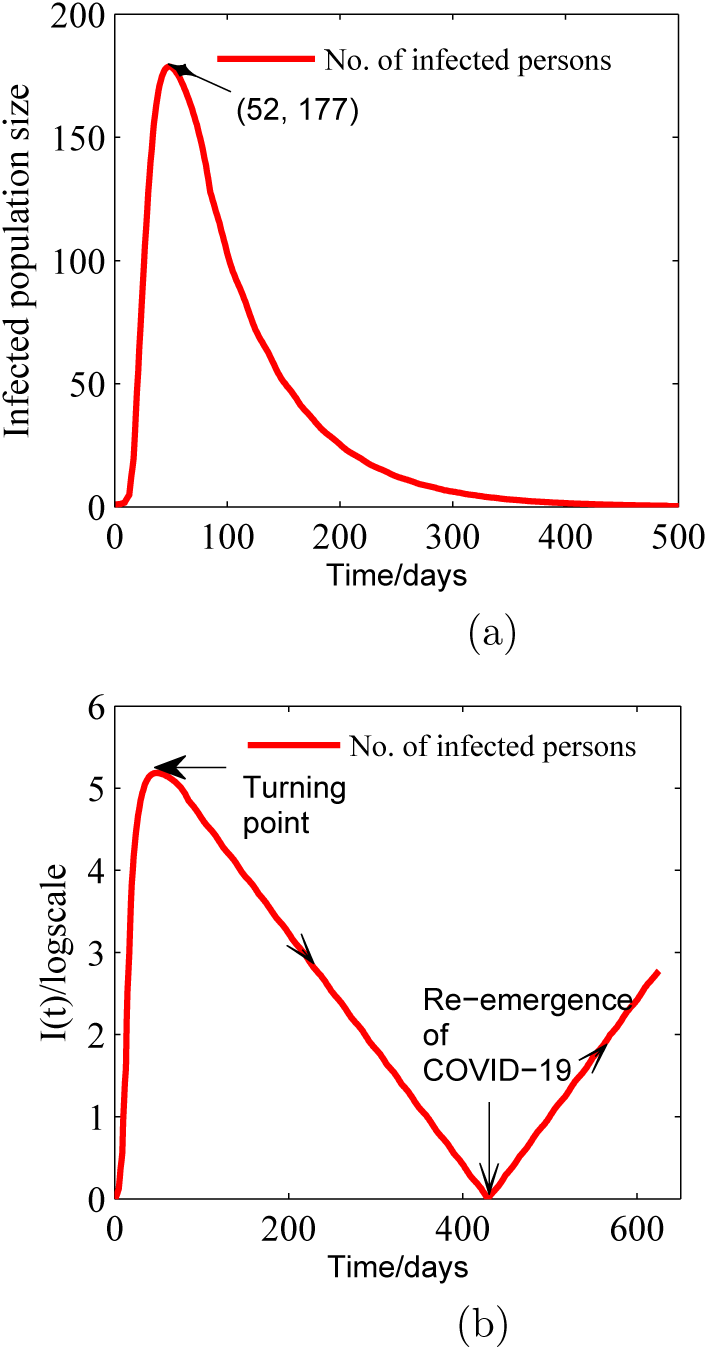
(a) The number of infected persons as a function of time to linear scale. (b) The number of infected persons as a function of time to logarithm scale for *R_n_*_0_ = 0.46

Fig 7 (a) shows the effect of varying the rate *ζ* at which awareness programs are implemented on the unaware population. An increase from *ζ* = 0.345 to *ζ* = 3.45 leads to a reduction in the number of the unaware persons. This means that the population is able to avoid the transmission of the disease. A decrease from *ζ =* 0.345 to 3.45 leaves a proportion of individuals unaware which means the authorities would need more resources to disseminate the information. Moreover, Fig 7 (b) explains the effect of varying of media *M* that disseminate information about COVID–19. We observe that reducing *M* from 4 to 2 leaves a high number of the population unaware of the existing measures. Whereas increasing M from 4 to 6 reduces the number of unaware sub–population implying people can be able to change the way they socialize; thus reducing the transmission of the disease. From Fig 7, it would take policy makers 400 days (≈ 1 yr and 10 days) from the inception of the disease to implement the awareness program and cover the entire population and realize stability. To prevent recurrence of disease in the population, continuous reminders need to continue because retention of information by individuals fades and individual behaviors towards prevention and control measures change overtime.

**Fig. 7.**
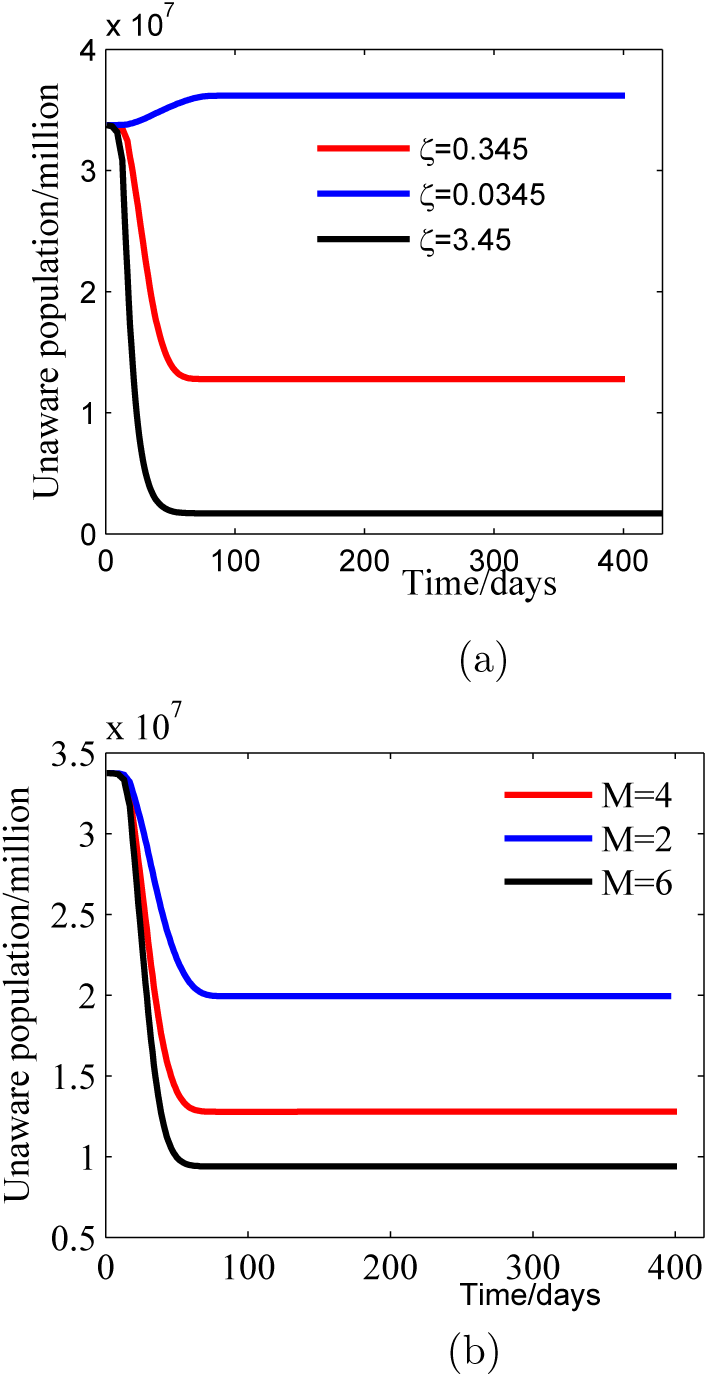
(a) Effect of varying the rate at which awareness programs are implemented on the unaware population. (b) Effect of varying the number of awareness programs on the unaware population.

## Conclusion

Regarding COVID–19 pandemic situation in Uganda, we proposed an SEIR epidemic model that incorporated prevention and intervention measures. This research illustrates capabilities of the SEIR model in predicting and therefore informing the general public about the impact of COVID–19 using a mathematical approach. The results obtained will be used to predict, inform and monitor the progress, timing and magnitude of the COVID–19 pandemic in Uganda.

We computed the reproductive number and it worked out as *R_n_*_0_ = 0.468. We note that *R_n_*_0_ is less than unity, thus forecast that several strategies in combination (including travel restrictions, mass media awareness, community buy–in and medical health interventions) will eliminate the disease from the population. However, our model predicts a recurrence of the disease after one year and two months (430 days) thus the population has to be mindful and continuously practice the prevention and control measures.

There is need for collaborative effort from citizens especially truck drivers and neighbours from Eastern African region in order to combat COVID–19 pandemic. In addition, there should be focus on strict inland mediation and awareness at borders including nearby villages in order to reduce on exogenous imported cases.

It is important to ensure fast detection, awareness, treatment and sufficient medical supplies are maintained. It is also important that persons with mild symptoms are maintained in institutional facilities.

We recommend that the Sub–Saharan countries including East Africa should adopt the model used to construct reliable intervention strategies to eliminate COVID–19 pandemic.

The question why Severe Acute Respiratory Syndrome Corona Virus 2 (SARS–CoV–2) re–emerges after 1 year and 2 months, shall be answered by the model we intend to embark on soon.

The research team intends to further conduct empirical studies in our local communities in order to inform the public about the impact of COVID–19 especially on prevention and intervention measures in Uganda. The stigma faced by recovered persons calls for special attention. There is need to inform and sensitize the community on how to cope and live with the victims.

## Data Availability

All relevant data used in the manuscript has been cited

## Conflicts of Interest

The authors declare no conflicts of interest.

## Funding

The research group is not funded. The team selflessly voluntarily worked to warrant that results are obtained in real–time, to monitor the progress of the COVID–19 pandemic.

## Author contributions

**Conceptualization**: Fulgensia Kamugisha Mbabazi, Awichi Richard, Gavamukulya Yahaya.

**Methodology**: Livingstone S. Luboobi, Fulgensia Kamugisha Mbabazi.

**Qualitative and numerical analysis of the model**: Fulgensia Kamugisha Mbabazi.

**Writing original draft–editing**: Fulgensia Kamugisha Mbabazi, Peter Olupot Oluput, Gavamukulya Yahaya, Saphina Biira, Samson Rwahwire.

**Writing final Manuscript**: Fulgensia Kamugisha.

**Review & editing**: Livingstone S. Luboobi, Awichi Richard, Saphina Biira, Gavamukulya Yahaya.

## Notes

### Competing Interest Statement

The authors have declared no competing interest.

### Funding Statement

No exteternal funding was received

